# Supporting Respiratory Epithelia and Lowering Inflammation to Effectively Treat Common Cold Symptoms: A Randomized Controlled Trial

**DOI:** 10.1101/2024.03.27.24304989

**Authors:** Pavel Pugach, Nazlie Sadeghi-Latefi

## Abstract

Common cold viruses are leading triggers of asthma attacks, causing nearly two million hospitalizations per year and productivity losses approaching $40B. They also increase susceptibility to bacterial infections driving antibiotic use. Post-market clinical studies have questioned the efficacy of most over the counter (OTC) cough and cold ingredients against placebo in treating various symptoms. To our knowledge, only aspirin significantly improved overall illness severity compared to placebo and that was by about 25-30%. In this double-blind randomized placebo-controlled trial involving 157 participants, we sought to determine whether a throat spray containing a mucosal immune complex (MIC) (comprised of lysozyme, lactoferrin, and aloe) can increase the hereto reported efficacy of aspirin at reducing common cold symptoms. Previously published reports showed that the MIC can protect respiratory epithelia and lower inflammatory cytokines. Participants self-administered treatments (throat sprays every hour and tablets every four hours) and completed surveys at home over two days. Treatments included MIC spray mixed with 6mg aspirin + placebo tablet (Treatment 1), MIC spray + placebo tablet (Treatment 2), MIC spray + 325 mg aspirin tablet (Treatment 3). Participants included adult volunteers ages 21-66 (average 44), 54% female, 46% male, 46% African American, 8% Asian, 39% Caucasian, and 7% Hispanic, having common cold symptoms lasting less than two days. The main outcome measures included Sore Throat Pain Intensity (STPIS) 0-100 at 36 hours (primary endpoint) and Modified Jackson Score (MJS), a combination of eight cold symptoms (secondary endpoint).

Both primary and secondary endpoints were met. Sore throat pain as measured by STPIS decreased 68-75% by 36 hours depending on treatment. Other symptoms such as nasal discharge, congestion, sneezing, cough, sore throat, and malaise as measured by MJS decreased 38-68% depending on treatment. In repeated measure within group analysis observing the same participants over multiple time points; STPIS mean change from baseline to 36 hours was as follows: Placebo (-7.84 (-14%) [95% CI -14.20 to -1.47]; p<0.0001), Treatment 1 (-42.41 (-75%)[95% CI -48.30 to -36.52]; p<0.0001), Treatment 2 (-38.60 (-68%)[95% CI -46.64 to -31.56]; p<0.0001), and Treatment 3 (-44.19 (-79%) [95% CI -52.11to -36.27]; p<0.0001). In repeated measure within group analysis all treatments significantly reduced cold symptom severity (MJS) from Days 1-2. Results were as follows: Treatment 1 (-2.26 (-38%) [95% CI -3.04 --1.47] p<0.0001), Treatment 2 (-3.81 (-53%) [95% CI -4.82 - -2.80] p<0.0001), Treatment 3 (-4.49 (-69%) [95% CI -5.62- -3.57]; p<0.0001).

As a result of this study, we conclude that supporting upper respiratory epithelia and reducing COX-mediated inflammation may be used to effectively treat common cold symptoms.

Trial registration ClinicalTrials.gov Identifier: NCT06106880Posted 30/10/2023

## INTRODUCTION

The common cold is a symptom-based disease caused by regularly circulating respiratory viruses excluding influenza and SARS or MERS associated coronaviruses (1) . Its economic impact is estimated at $40 billion in the US (2, 3). Despite the public health burden, there are no clinically proven, FDA approved drugs, or other remedies to effectively lower symptom severity or shorten duration of illness(1).

Over the counter (OTC) drugs for common cold symptoms contain ingredients allowed by the FDA under OTC Monographs and their administrative orders as of 1972. Since then, post-market clinical studies have questioned the efficacy of many of these ingredients against placebo in treating various symptoms. Efficacy of dextromethorphan (4-8), guaifenesin (9, 10), pseudoephedrine (11) and benzocaine (12, 13) have all been questioned. The efficacy of pseudoephedrine for the treatment of nasal congestion is questionable ^(14, 15)^, and it has been shown to exacerbate conditions such as hypertension and restless leg syndrome (11, 16). Its role as a key ingredient in the formulation of illicit substances led to its behind-the-counter regulation(17) and has subsequently been replaced by phenylephrine in OTC cold products over the last 15 years. Several studies found that phenylephrine is not different from placebo in treating cold symptoms (18-22). In September 2023, as FDA panel issued the ruling that oral phenylephrine, grossing over $1.5 billion in the last year alone, is not effective for the treatment of cold and flu symptoms (23). Ibuprofen, and acetaminophen effectively improve mainly fever and pain symptoms (24, 25); but to our knowledge, not any validated measure of overall illness. Furthermore, according to some studies, prenatal use of acetaminophen has been associated with a 19% and 21% increase in the risk of autism spectrum disorder and attention deficit disorder, respectively (26). In addition, therapeutic doses of acetaminophen have been shown to alter liver function, as well as significantly deplete glutathione, an important endogenous antioxidant (27-30). To our knowledge, only aspirin (with vitamin C) significantly improved overall illness severity as measured by the Wisconsin Upper Respiratory Symptom Score by about 25-30% (31). Aspirin is a well-known irreversible COX-enzyme inhibitor. COX enzymes have been shown in numerous studies to induce prostaglandin formation which leads to common cold symptoms (32-36). Upon sensing injury at the respiratory epithelium, bradykinin induces release of arachidonic acid (AA) from cell membranes via phospholipase A2, and AA is then converted to prostaglandin E2 via COX enzymes (37). Repairing the epithelia and controlling inflammation are critical to limiting symptoms (38).

We treated people exhibiting naturally acquired common cold symptoms with a throat spray containing a Mucosal Immune Complex (MIC) and various combinations of aspirin, wintergreen oil, and menthol. The aspirin was either mixed into the throat spray (6 mg) or taken as a tablet (325mg). The MIC contained lysozyme, lactoferrin and aloe, natural dietary supplements which lubricate and protect the respiratory barrier (39) and which may also affect rheological properties of the mucosal surface (40) or act as non-specific glycoprotein attachment inhibitors (41). Lactoferrin binds to multiple viruses, blocking their entry into epithelial cells, induces type I interferon production and enhances Th1 responses in the context of viral infection (42-46). Lactoferrin also prevents and repairs the virus-induced cytotoxicity in host cells, thereby limiting the release of damage-induced pro-inflammatory cytokines that correlate with symptoms (46). Lysozyme has antimicrobial effects and may work in synergy with lactoferrin through unknown mechanisms (47). Reduced levels of lysozyme and lactoferrin in the mucosa increases susceptibility to infections and leads to more severe illness, further supporting their role in mucosal health (48, 49). According to previously published studies using human respiratory organoid tissues, the MIC augmented aspirin’s anti-inflammatory effects possibly by protecting or buffering the respiratory epithelia(39).

## METHODS

The protocol for this randomized, placebo-controlled clinical trial was approved by the Advarra Institutional Review Board and written informed consent was obtained for all participants. This study followed the Consolidated Standards of Reporting Trials (CONSORT) reporting guidelines. The protocol for this active-comparator, parallel-arm RCT (NCT06106880) has been published on clinicaltrials.gov.

### Trial Participants

The trial was an 11-month multi-center randomized clinical trial conducted from May 2022 to June 2023 in participants’ homes in Washington DC, Baltimore MD, New York NY, Atlanta, Georgia, Houston TX, and Orange County CA. Inclusion criteria were: healthy adults aged 18-65, experiencing a sore throat rated at least 3 on a 10-point scale, and a sore throat duration of less than 48 hours at the time of intake assessment. Exclusion criteria included: sore throat exceeding two full days, likelihood of strep throat, allergies to eggs, milk, or aspirin, pregnancy, presence of chronic disease, recent history of allergy, fever above 101°F, ACE inhibitor use, participation in another clinical trial, and smoking.

Subjects were recruited through targeted social media advertisements. Interested individuals contacted the Clinical Trial Manager (CTM) who conducted a phone interview using a standardized script. Responses were recorded in a digital form (CTM Intake Form) on a HIPAA-compliant Survey Monkey™ platform. Participants deemed eligible were randomized, seen at home by the Clinical Trial Administrator (CTA), consented, and given blind coded kits containing treatment or placebo. Prior to commencing treatment the following morning, participants met with the PI by video conference.Personally identifiable information was securely stored on password-protected mobile devices and databases, accessible only to authorized personnel.

### Randomization and Blinding

180 eligible participants were randomized 1:1:1:1 to receive one of three treatments or placebo. Kits containing treatments or placebo, instructions, and survey questionnaires were provided to patients at home after signing informed consent. Randomization was performed by a third party CRO who manufactured and bottled the treatments and placebo. Codes were generated by a computer program using the rand function which uses the Mersenne Twister algorithm. A total of 400 codes were assigned, with each code representing one kit, then four treatment groups were established for participant allocation. Random codes were generated in blocks of 8 to maintain balanced group sizes after every 8 patients were randomized. The code format consisted of a two-digit investigator number followed by a four-digit subject identifier (e.g., XX-XXXX), enabling unique identification of each participant and their assigned kit. Throughout the study and until statistical analysis was complete only the CRO and the physician on call for adverse events, the Adverse Events Specialist (AES) was informed which kit numbers were assigned to which treatment groups.

### Intervention Groups

The four intervention groups included: MIC spray containing 0.6% aspirin (6mg per spray dose) + placebo tablet (Treatment 1), MIC spray + placebo tablet (Treatment 2), MIC spray + 325 mg aspirin tablet (Treatment 3). The MIC was composed of 0.5% bovine lactoferrin, 5% lysozyme, and 0.2% whole leaf aloe vera juice. Treatments 1, 2, and 3 sprays contained menthol at a concentration of 0.5% (5mg per spray dose), and Treatments 2 and 3 sprays also contained wintergreen oil at a concentration 0.6% (6mg per spray dose). The placebo spray contained 0.0009% (0.009 mg per spray dose) menthol, a sub-therapeutic dose. The throat sprays, aspirin pills, placebo pills and placebo throat sprays were identical in taste and appearance.

Sprays were administered each waking hour for two days. Sprays delivered 0.5mL per actuation from each side of a two-sided spray bottle for a total of 1ml per dose. One set of two sprays was recommended per waking hour for a total of 12mL per day or 72mg of aspirin. The pills were taken every 4 hours. A daily dose of 1.372g of aspirin was administered, which falls within the recommended adult daily dose of the US Food and Drug Administration.

Once enrolled, per the protocol, participants were seen via telemedicine by the Principal Investigator (PI). The PI directed them to begin treatment as well as a checklist of daily surveys on the morning of the following day. Immediately before their first treatment and at 4-hour intervals during waking hours (3 times per day), participants recorded their pain levels on the STPIS Visual Analog Scale (VAS). A Jackson score questionnaire was completed at the end of each day to assess symptom intensity.

Once the two days were completed, the participants mailed their forms to the PI. The PI and CTM were available to address non-urgent matters, while the Adverse Events Specialist (AES) handled serious adverse events and unblinded a participant if necessary.

### Outcomes Measures

The primary outcome was difference in pain intensity on the STPIS over a 36-hour period following the administration of the first dose of medication, as measured by the STPIS VAS on a 100mm scale. The application of visual analog scales for assessing sore throat pain, as well as various other types of pain, has been widely validated through extensive research studies (50). All STPIS lines on the paper patient forms were 100mm in length with “no pain” on the left side and “severe pain” on the right side of the line. Participants were instructed to physically mark the place on the line that corresponded to the pain they felt in their throat. The patient forms were then digitally scanned by Econometrica Inc Research and Management, Bethesda, Maryland, the data management company (DMC) as they were received. To convert the marks to numeric values, the DMC used the Adobe measuring tool to measure the length of the entire line and normalize for minor changes that may have resulted from the scanning process. This was calculated as the denominator. The numerator was then calculated as the distance from the left side of the line to the mark (or the center of the mark if the mark was not perpendicular). The division of the numerator by the denominator then gave the STPIS value which was converted to a 100 point scale (multiplied by 100) to generate the final STPIS value.

The secondary outcome was reduction in cold symptoms such as fever, sneezing, coughing, chills, and malaise by the second day, as measured by changes in the Modified Jackson Score, a well-validated measure of common cold symptoms (51, 52). To perform quantitative analysis of Jackson Scores, the categorical responses were recoded to numeric values, based on the following formula: Absent= 0, Mild= 1, Moderate= 2, Severe= 3. Next, the Modified Jackson Score was calculated as the sum of the numeric values from all symptom assessments. The total symptom score could range from 0-24 with higher numbers representing higher symptom burden.

### Data Acquisition

Participants mailed their survey forms to a secure PO Box accessible only to the PI. Forms did not contain any personal information, only the randomized kit number assigned to that participant. The PI did not share the data with anyone besides the DMC to which they were delivered and scanned for blinded analysis.

### Statistical Analysis

All statistical analyses were carried out blind by the DMC. Treatment groups were unmasked by the AES when the data analysis was complete. The primary endpoint was change in STPIS from treatment initiation to 36 hours (Day 1, 1^st^ entry to Day 2, 4^th^ entry). The secondary endpoint was change in Modified Jackson Score (MJS) from Day 1 to Day 2. The mean, 95% confidence interval (CI) of the mean, standard deviation, and median were calculated for each measurement (for both days) to demonstrate the basic characteristics of the data.

To analyze the changes in scores over time and assess the impact of the interventions, repeated-measures ANOVA were used. In addition, post-hoc analysis – pairwise group comparisons – was conducted to identify specific differences between each group pair after finding a statistically significant overall impact.

## Sample Size Justification and Interim Analysis

A previous study utilizing an STPIS endpoint (50) following administration of flurbiprofen (an NSAID) reported a 59% decrease (-196.6mm x h [95% CI -321 to -72.2]; p<0.01) using as its endpoint a time weighted sum difference over 24 hours. A total of 198 subjects was the sample size for this two-group RCT with 1:1 randomization that compared placebo to treatment (50). This same sample size was adopted for our study.

In our placebo controlled RCT with three active treatment arms using various doses and 1:1:1:1 randomization, the independent DMC conducted an interim analysis based on 159 subjects. Efficacy of all three active treatment groups far exceeded expectations – a highly statistically significant difference was seen among all treatment groups when the primary endpoint was examined (p < 0.0001). The three active treatments showed on average a 70% improvement in the primary efficacy endpoint relative to placebo. Analyses of secondary endpoints as well as between-group comparisons were performed and reviewed by the DMC before the treatment assignments were unblinded.

Analysis of secondary endpoints showed similar levels of improvement relative to placebo. Therefore, with 40% of the projected sample size accrued and analyzed, the decision was made to terminate the study early based on 159 subjects (approximately 40 subjects per group).

## RESULTS

### Participants, Retention, and Fidelity

Of 350 individuals initially screened by phone (by the CTM), 180 were subsequently screened in person (by the CTA), consented, and randomized (Fig. 1). Following randomization, 23 individuals were lost to follow up: 1 individual missed their appointment with the PI, 1 was discontinued for an adverse event, 3 were disqualified by the PI, 16 did not mail in their patient forms and were unreachable, and 2 returned illegible patient forms as determined by DMC before unblinding. Randomized individuals were similar to those lost to follow up as seen in Table 1. Of those randomized, 44 were allocated to placebo, 49 to Treatment 1, 44 to Treatment 2, and 43 to Treatment 3. All participants were analyzed in their randomized group, and there was no between arm cross-over. In the final analyses there were 35 in the placebo group, 43 in Treatment 1, 42 in Treatment 2, and 37 in Treatment 3 due to loss to follow up as explained above and as seen in Figure 1. The one adverse event was deemed unrelated to treatment by the AES (Fig. 1 and Table 1).

**Table 1.** Demographics of participants randomized, deemed ineligible, refusing to participate, and lost to follow up.

| Variable | Randomized<br>(n=180) | Ineligible<sup>b</sup> (did not meet inclusion or<br>met exclusion criteria)<br>(n=154)<br>n=134 met exclusion criteria +<br>n=20 missed CTA visit | Refused<sup>b</sup><br>(n=16)<br>n=1 admin error <sup>c</sup> +<br>n=15 declined<br>participation | Lost to Follow Up<br>(n=23)<br>n=16 forms not received +<br>n=2 forms illegible +<br>n=1 discontinued +<br>n=3 PI disqualifications +<br>n=1 missed PI appt |
| --- | --- | --- | --- | --- |
| Mean age (SD) | 43.5 (13.5) | NA | 40.7 (12.8) | 38.7 (12.4) |
| BMI (SD) | 27.4 (6.2) | NA | NA | 26.3 (6.4) |
| Male Gender (%) | 46.6 | NA | 43.8 | 45.5 |
| Atlanta, GA site (%)<sup>a</sup> | 35.6 | 9.5 | 6.4 | 25 |
| Mean Intake Sore Throat Rating (0-10) (SD) | 5.9 (1.6) | 4.2 (2.1) | 5.7 (1.6) | 5.1 (1.4) |
| Caucasian (%) | 39.4 | NA | NA | 40.9 |
| African American (%) | 43.9 | NA | NA | 27.3 |
| Hispanic/Latinx (%) | 10 | NA | NA | 31.8 |
| Asian (%) | 6.7 | NA | NA | 0 |
| Sore throat duration at intake (in hours) | 30.2 | NA | 31.3 | 35.8 |
| Mean body temperature (°F) | 97.0 | NA | 98.4 | 98.8 |

<sup>a</sup> Of 6 US study sites, Atlanta, GA had the greatest enrollment, followed by New York City at 27.7%.
<sup>b</sup> NA, data was not collected for these participants because they were deemed ineligible before the data collection step of the protocol.
<sup>c</sup> PI missed appointment, administrative error

**Fig 1.**
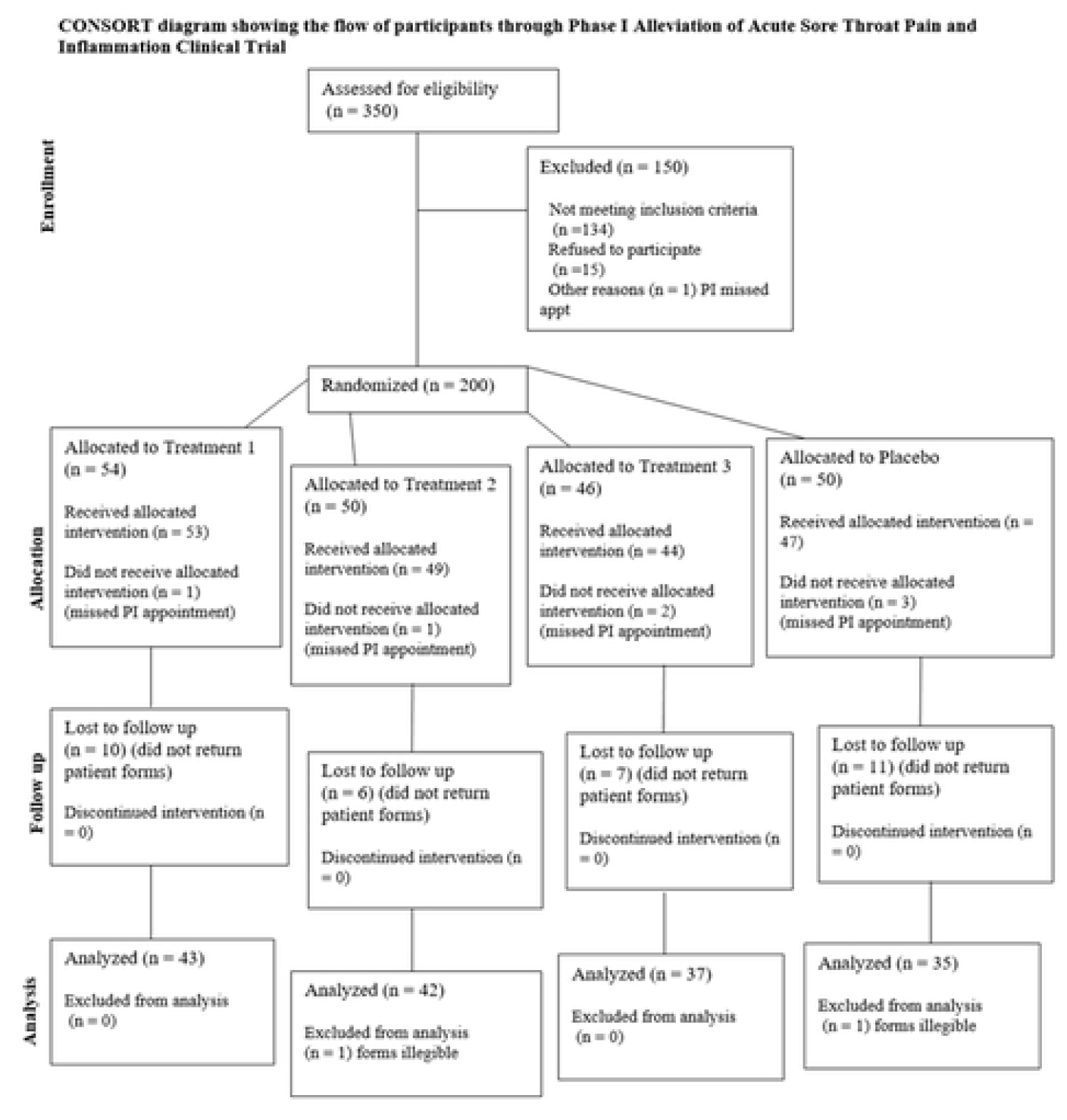
Participant Flow Diagram. CONSORT diagram showing the number of participants who were assessed for eligibility, excluded, randomized, and lost to follow up through the course of the study.

Per Table 1, the 180 randomized participants had baseline mean (SD) age of 43.5(13.5) years, and BMI of 27.4 (6.2). A total of 96 (52.5%) participants were women; 79 (43.9%) were African American, 18 (10%) were Hispanic/Latinx, 71 (39.4%) were Caucasian, and 12 (6.7%) were Asian. The attrition rate (participants lost to follow up) was 12.8% which included the 23 participants lost to follow up.

All tables and graphic representations of data were prepared by the DMC.

### Primary Outcome

We used STPIS to evaluate the Treatment effects on sore throat pain over time. Effects of STPIS over time was analyzed using repeated measures analysis to measure within subject changes. The mean, 95% confidence interval (CI) of the mean, standard deviation, and median for STPIS measures (Fig. 2a and 2b) summarize the distribution and central tendency of the data.

**Fig 2.**
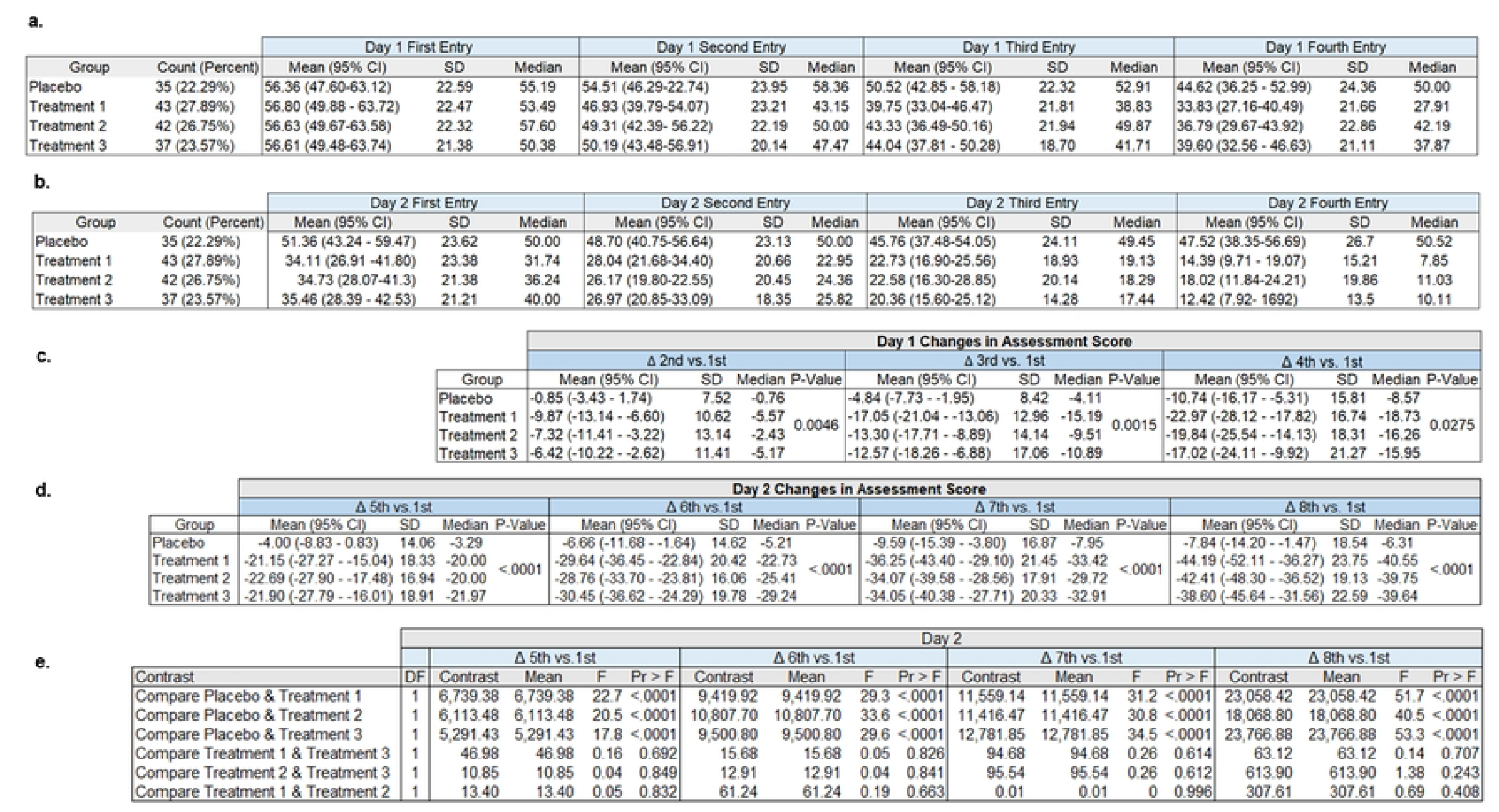
Assessment of STPIS scores. Tables show calculation of average STPIS scores across groups and results of statistical analyses. STPIS analyses were obtained four times per day over two days for each participant. Descriptive statistics for Day 1 (a), Day 2 (b), changes in STPIS assessments between Day 1 and 2 (c), and pairwise comparisons of STPIS changes between Day 1 and 2 are shown (d). Change in STPIS score from the fourth measure of the second day compared to the first meausure on the first day was the primary endpoint of the trial.

STPIS Day 2, 4^th^ entry (36 hours after treatment initiation), was statistically significant from baseline (1^st^ entry) for each of the treatments (p< 0.0001) as well as placebo (Fig. 2b). The mean changes were placebo (-7.84 [95% CI -14.20 to -1.47]; p<0.0001) (-14%), Treatment 1 (-42.41 [95% CI -48.30 to - 36.52]; p<0.0001) (-68%), Treatment 2 (-38.60 [95% CI -46.64 to -31.56]; p<0.0001) (-75%), and Treatment 3 (-44.19 [95% CI -52.11to -36.27]; p<0.0001) ((Fig. 2d and Fig. 3). Additional post hoc analyses (pairwise comparisons) found statistically significant differences between placebo and each of the treatments (p< 0.0001) (Fig. 2e) but not between the treatments themselves. The changes on Day 2 were greater than for those on Day 1 and the significance of the differences between the groups was greater on Day 2 than on Day 1(Fig. 2c and 2d).

**Fig 3.**
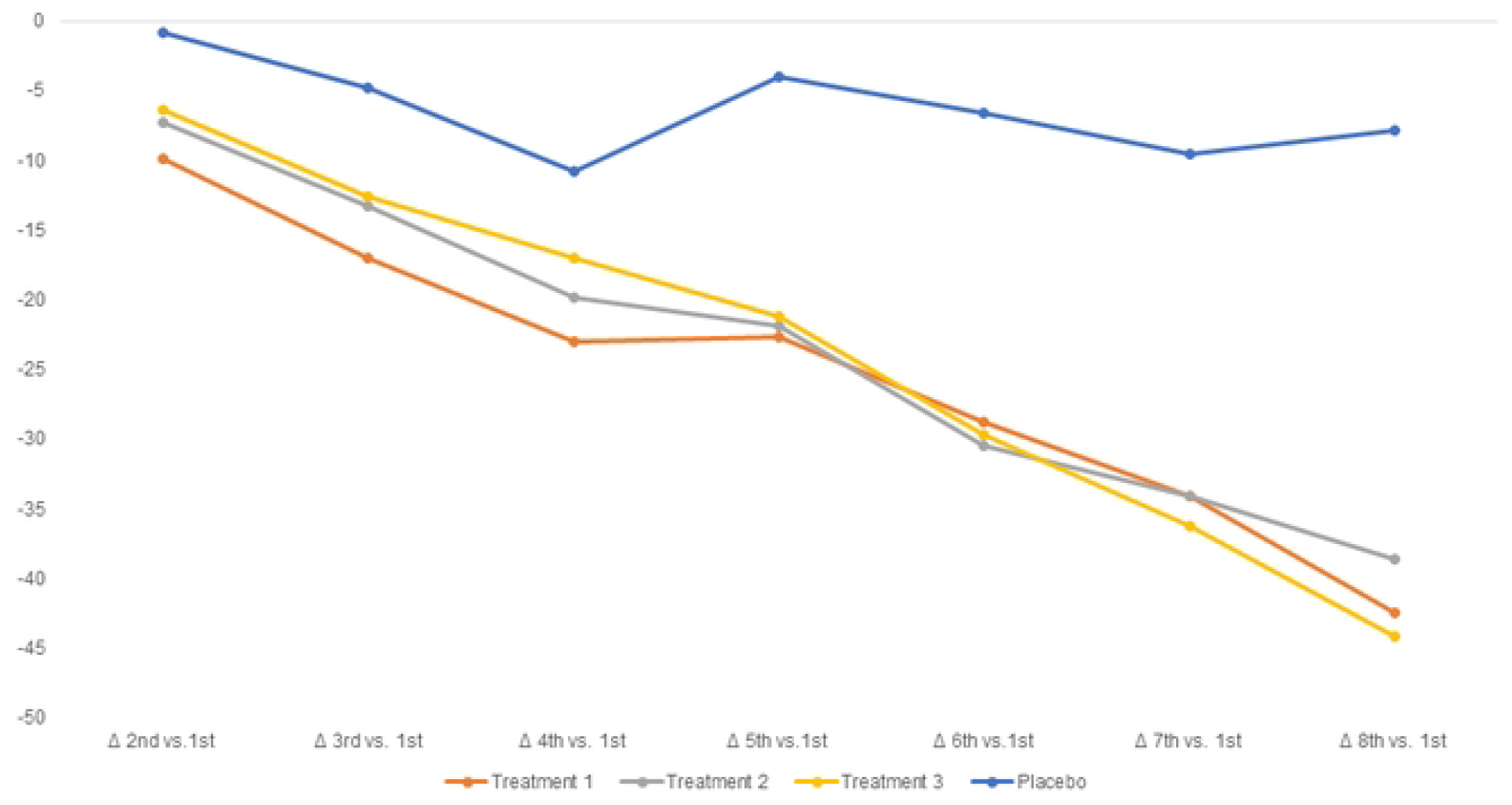
Changes in the STPIS means by group. Changes in the mean STPIS score at the first assessment on Day 1 to the last assessment on Day 2 (the primary endpoint) are shown for each treatment group. Numerical values of those graphed can be seen in Fig. 2d.

### Secondary Outcomes

Effects on the eight sets of Jackson Score measures were analyzed using repeated measures analysis to measure within subject changes on MJS. MJS includes a symptom severity questionnaire for eight symptoms: sneezing, nasal discharge, nasal congestion, sore/scratchy throat, cough, headache, malaise and fever/chills. Each is from 0 to 3 (0=absent, 1=mild, 2=moderate, 3=severe) (51). Significant differences on MJS were found between Treatments on Day 2 (p < 0.0001) (Fig. 4a,b,c). Treatment 3 exhibited the largest mean change, indicating the most significant improvement in symptom severity compared to the other groups (Fig. 4a,b,c) (-4.59 [95% CI -5.62 to -3.57]; p<0.0001). Post hoc pairwise comparisons revealed significant differences between Treatment 1 and Treatment 3, as well as between Treatment 1 and Treatment 2, each treatment and placebo (p<0.0001) (Fig. 4c). Individually, each of the eight symptom scores showed some significant differences between groups on Day 2.

**Fig 4.**
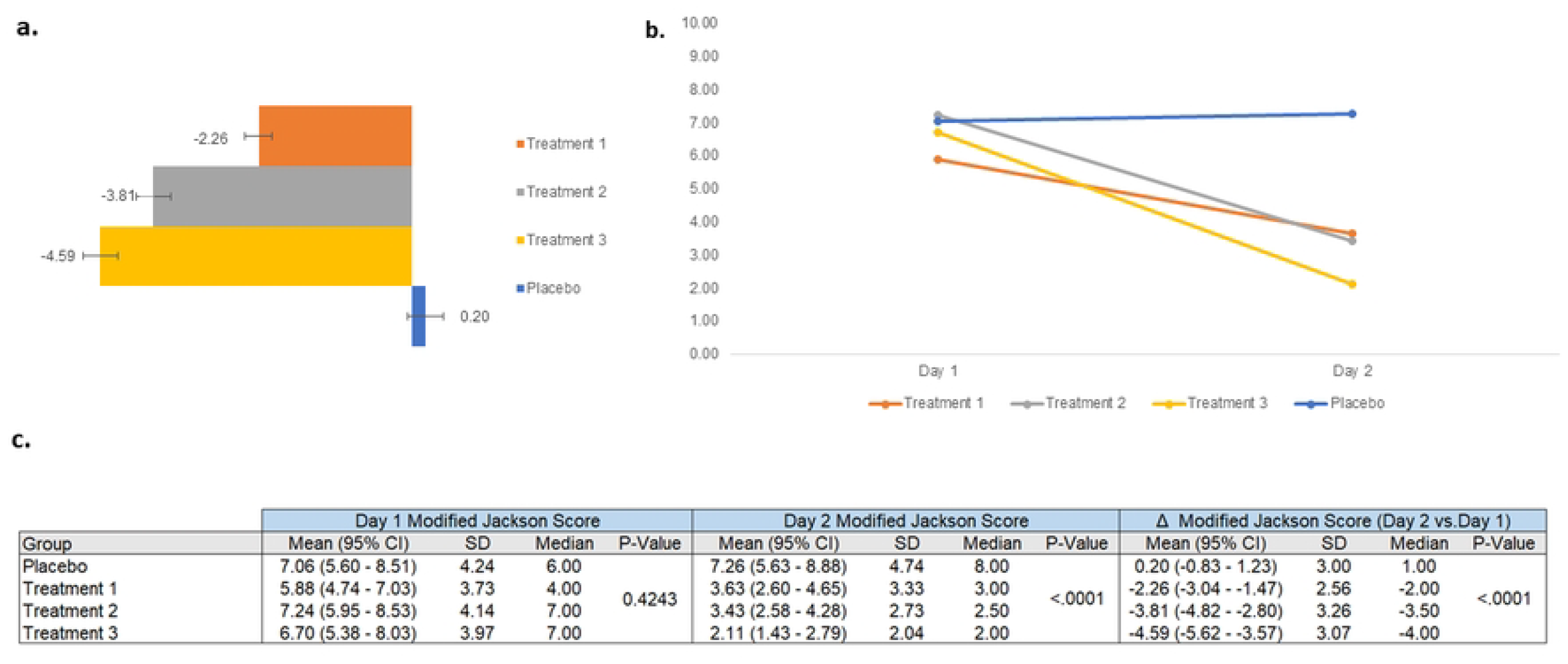
Assessment of Jackson Scores. Mean Jackson scores on Day 1 and Day 2 are shown for each treatment group in a bar graph along with the numerical value for the mean and an error bar depicting the standard error of the mean (a). The same values are also shown in a line graph (b). Statistical analyses are also shown (c).

For nasal congestion, Treatments 2 and 3 were significantly better than placebo and Treatment 1 but not different from each other (Fig. 5a). From Day 1 to Day 2 the mean score for Treatment 1 declined by 0.26 points (27.6%) compared to 0.06 points (5.5%) for placebo. For Treatments 2 and 3 nasal congestion decreased by 0.69 points (57%) and 0.71 points (66.7%) from Day 1-2.

**Fig 5.**
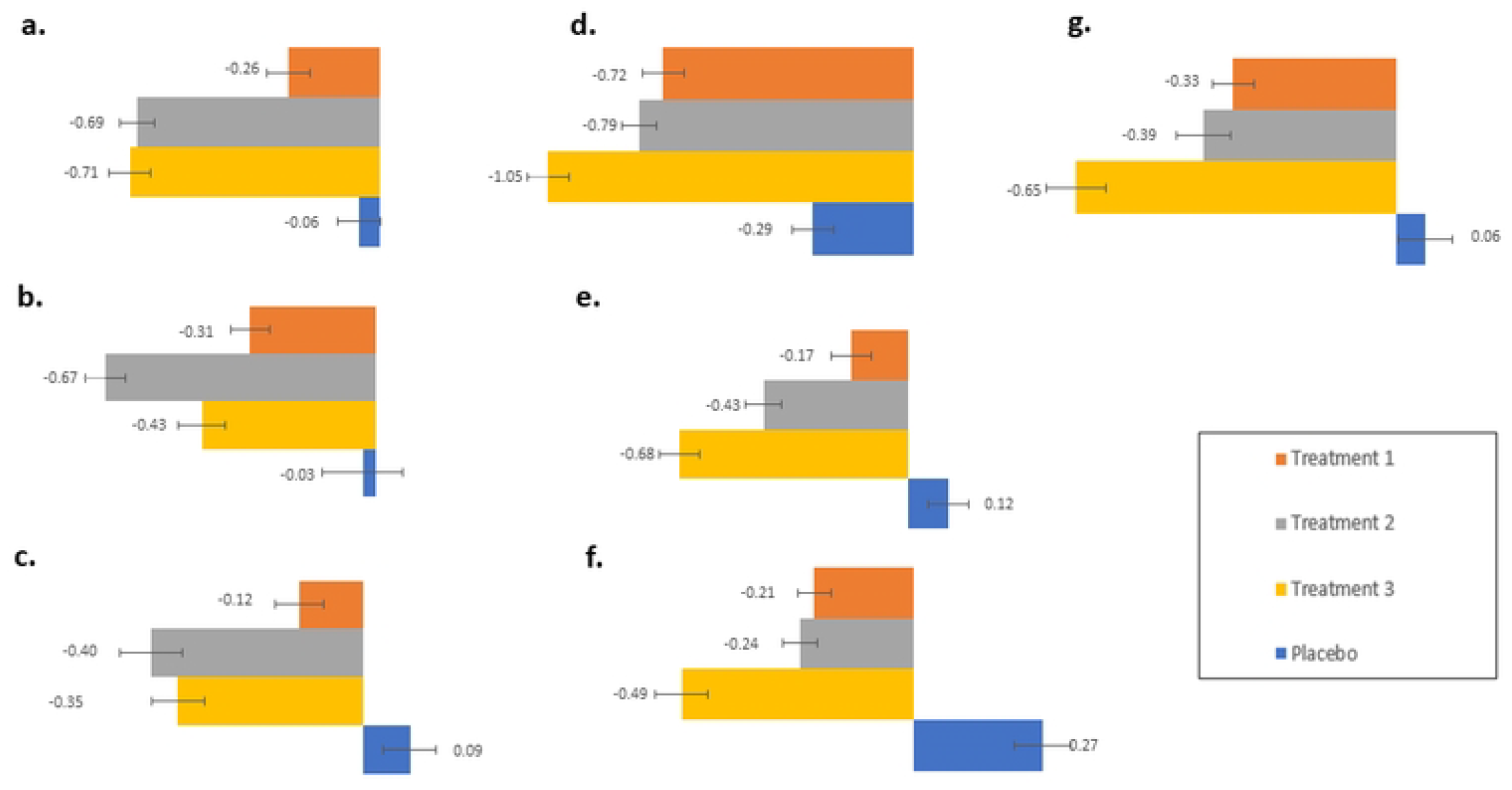
Assessment of individual symptoms. Changes in mean scores for individual symptoms from Day 2 to Day 1 for each treatment group are shown in a bar graph along with numerical values and an error bar depicting standard error of the mean: nasal congestion (a), nasal discharge (b), sneezing (c), sore throat (d), cough (e), headache (f), and malaise (g).

For nasal discharge, Treatment 2 produced the strongest effect (Fig. 5b). The effect of Treatment 2 was statistically better than placebo and also Treatment 1. The effects of Treatment 3 compared to placebo were also significant albeit showed less decrease than Treatment 2. The differences between Treatments 2 and 3 were not statistically significant. Treatments 1-3 decreased nasal discharge scores by 0.31 (42.1%), 0.67 (59.2%), and 0.43 (58.1%) respectively.

For sneezing, Treatments 2 and 3 produced the strongest effects (Fig. 5c). The effect of Treatment 2 and 3 were statistically better than placebo. The differences between Treatments 2 and 3 were not statistically significant. Treatments 1-3 decreased sneezing scores by 0.12 (34%), 0.4 (47.7%), and 0.35 (56.5%) respectively. The effects of Treatment 1 were not significantly different from that of placebo for any of the nasal symptoms. Both Treatments 2 and 3 used throat sprays containing wintergreen oil which may have contributed to decreasing all nasal symptoms, however aspirin did not seem to provide any additional benefit on this measure.

Participants in the placebo group reported a decrease of 17.5% in sore throat scores between Day 2 and Day 1. Nevertheless, all treatments showed statistically significant decreases compared to placebo with Treatment 3 showing the largest decrease (Fig. 5d). The effect of Treatment 3 was statistically different from Treatment 1 but not Treatment 2. Treatments 1-3 decreased sore throat scores by 0.72 (45%), 0.79 (53.5%), and 1.05 (66%) respectively. The results of the sore throat Jackson Score measure showed similar trends as seen with STPIS, however differences among the groups were not statistically different for STPIS. This may be explained by the STPIS measuring changes from the initiation of treatment (morning of Day 1) whereas the Jackson Score measured change from Day 1 (end of day) to Day 2 (end of day), or that the STPIS included more frequent measures (12 times a day versus 1 time a day).

Treatment 3 had the strongest effect on cough (Fig. 5e). Treatment 3 was statistically different from placebo and Treatment 1, but not from Treatment 2. Treatment 2 also showed statistically significant improvement compared to placebo. Treatment 1 was not statistically significant from placebo (Fig. 5d). Treatments 1-3 decreased cough scores by 0.17 (23.4%), 0.43 (46.2%), and 0.68 (71.6%) respectively. Both Treatments 2 and 3 used throat sprays containing wintergreen oil which may have contributed to decreasing cough symptoms. Aspirin may provide some additional benefit on this measure.

All treatments showed a statistically significant improvement in headache and malaise scores compared to placebo (Fig. 5f and 5g). Treatment 3 had the strongest effect and was significantly better than Treatment 1 but not Treatment 2. (Fig. 5f and 5g), Treatments 1-3 decreased headache by 0.21 (44.7%) and 0.33 (48.4%), 0.24 (48%) and malaise by 0.39 (50%), 0.49 (77.4%) and 0.65 (77.4%) respectively.

## DISCUSSION

The present study successfully met its primary and secondary endpoints. All Treatments showed significant improvements over placebo in treating common cold symptoms, more specifically, STPIS at 36 hours and MJS at 48 hours. STPIS began to improve upon the first treatment. By 36 hours, STPIS decreased 68-79% depending on treatment. The MJS which is comprised of eight symptoms, decreased 38% for Treatment 1, 52.6% for Treatment 2, and 68% for Treatment 3 between the first and second day after treatment initiation. On between group comparison for MJS from Day 1 to Day 2, Treatments 2 and 3 performed significantly better than Treatment 1.

This is the first study to investigate a treatment acting on the upper respiratory mucosa combined with systemic aspirin for treating upper respiratory cold symptoms. Aspirin on its own has been studied previously for treating common cold symptoms, but its effects were not as strong as those for our throat spray paired with aspirin (Treatment 3) or the spray alone (Treatment 2). A previous study of 800 mg aspirin paired with vitamin C showed a decrease in Wisconsin Upper Respiratory Symptom Survey Domain 2, a validated scale of common cold symptoms of 29% compared to placebo 2 hours after treatment initiation, a decrease of 30.2% at the end of the first day of treatment, and a decrease of only 12% by the second day^31^. Effects on Days 3 and 4 were not statistically different from placebo^31^. In another study where patients were given 800mg of aspirin, and assessed for 6 hours after treatment, sore throat pain intensity differences decreased 58% 2 hours after treatment(53). In the same study, 6 hours after treatment, headache was reduced 38% compared to 16% for placebo, muscle aches and pains were reduced 38% following treatment and 25% following placebo. Differences in sinus pain and fever were not different from placebo at 6 hours post treatment(53).

Even Treatment 1, the least effective treatment containing 6 mg of aspirin, decreased common cold symptoms more than previously observed with 800mg aspirin. Unexpectedly, an equivalent amount (6mg) of wintergreen oil (Treatment 2) in lieu of aspirin led to a greater improvement in nasal symptoms, but not in pain-associated symptoms (Fig.5). The addition of an aspirin pill to the wintergreen oil (Treatment 3) did not further improve nasal symptoms, but did improve pain-associated symptoms (Fig.5). This demonstrates that wintergreen oil offers an additional benefit for nasal symptoms, that commonly arise during upper respiratory tract infections. The wintergreen oil in Treatment 2 could be acting by several mechanisms and may explain the stronger efficacy of Treatment 2. In addition to methyl salicylate, wintergreen contains numerous essential oils, a mix of aldehydes, esters, ketones, peroxides, and phenols that can be inhaled and carried throughout the respiratory tract. These have been shown to have antimicrobial and anti-inflammatory effects(54). They may act on TRP ion channels in the airways which play a role in respiratory symptoms(54). Menthol also acts on TRP ion channels and is composed of a unique mix of terpenoids to wintergreen oil. It may have additive or synergistic effects when combined with wintergreen oil(54). Formulations in this study were made with natural menthol and wintergreen oil. Chemical compositions may be influenced by environmental factors and isolation processes, which should be standardized in future studies. Here we compared effects of our treatments to previously published effects of aspirin. Future studies should include treatments with aspirin alone to more directly compare the effects of aspirin with or without MIC, and should also control for the effects of wintergreen oil and menthol.

## Data Availability Statement

Data from individual patients will not be made available because patients were not asked if they would be willing to share their data as part of this study. Data from individual patients was analyzed in a blinded and anonymous format by the Data Management Company (DMC), a third party. Individual data can be made available upon reasonable request to the DMC to verify the results of this study.

